# Acute pharmacodynamic responses to exenatide: Drug-induced increases in insulin secretion and glucose effectiveness

**DOI:** 10.1101/2023.03.15.23287166

**Authors:** Simeon I. Taylor, May E. Montasser, Ashley H. Yuen, Hubert Fan, Zhinoosossadat Shahidzadeh Yazdi, Hilary B. Whitlatch, Braxton D. Mitchell, Alan R. Shuldiner, Ranganath Muniyappa, Elizabeth A. Streeten, Amber L. Beitelshees

## Abstract

**Background:** GLP1R agonists provide multiple benefits to patients with type 2 diabetes – including improved glycemic control, weight loss, and decreased risk of major adverse cardiovascular events. Because drug responses vary among individuals, we initiated investigations to identify genetic variants associated with the magnitude of drug responses.

**Methods:** Exenatide (5 µg, sc) or saline (0.2 mL, sc) was administered to 62 healthy volunteers. Frequently sampled intravenous glucose tolerance tests were conducted to assess the impact of exenatide on insulin secretion and insulin action. This pilot study was designed as a crossover study in which participants received exenatide and saline in random order.

**Results:** Exenatide increased first phase insulin secretion 1.9-fold (p=1.9×10^-9^) and accelerated the rate of glucose disappearance 2.4-fold (p=2×10^-10^). Minimal model analysis demonstrated that exenatide increased glucose effectiveness (S_g_) by 32% (p=0.0008) but did not significantly affect insulin sensitivity (S_i_). The exenatide-induced increase in insulin secretion made the largest contribution to inter-individual variation in exenatide-induced acceleration of glucose disappearance while inter-individual variation in the drug effect on S_g_ contributed to a lesser extent (β=0.58 or 0.27, respectively).

**Conclusions:** This pilot study provides validation for the value of an FSIGT (including minimal model analysis) to provide primary data for our ongoing pharmacogenomic study of pharmacodynamic effects of semaglutide (NCT05071898). Three endpoints provide quantitative assessments of GLP1R agonists’ effects on glucose metabolism: first phase insulin secretion, glucose disappearance rates, and glucose effectiveness.

**Registration:** NCT02462421 (clinicaltrials.gov)

**Funding:** American Diabetes Association (1-16-ICTS-112); National Institute of Diabetes and Digestive and Kidney Disease (R01DK130238, T32DK098107, P30DK072488)

## 1 | Introduction

Head-to-head comparative effectiveness studies demonstrate that diabetes drugs are not strongly differentiated with respect to their effectiveness to decrease mean HbA1c ^1^. Nevertheless, individual patients vary widely in their responses to individual therapies. For example, semaglutide (1.0 mg/wk) decreased HbA1c by an average of 1.64% (SEM ≈ 0.05%; SD ≈ 0.95%) in patients with baseline HbA1c=8.17% ^2^. The observed magnitude of standard deviation suggests that some patients experienced little if any decrease in HbA1c while others experienced >2.5% HbA1c-lowering. This inter-individual variation may result from genetics, environment, or interplay between both factors. Pharmacogenetics has potential to guide treatment decisions, enabling clinicians to recommend optimal therapy for individual patients based on individualized predictors of responsiveness and susceptibility to adverse effects ^1^.

Pharmacogenomic studies have identified genetic variants associated with the magnitude of responses to several diabetes drugs, including metformin, sulfonylureas, and GLP1 receptor agonists ^3–7^. While some genetic variants alter a drug’s pharmacokinetics, other genetic variants alter pharmacodynamics by altering functions of proteins that mediate drug responses. Although genetic variation in pharmacokinetics may be less relevant for responses to an injectable peptide drug such as exenatide, genetic variants in the *GLP1R* gene have been reported to be associated with pharmacodynamic responses to GLP1 receptor agonists ^4–7^.

We initiated this clinical trial as a pilot and feasibility study for our ongoing genome-wide association study of responses to semaglutide (NCT05071898). Specifically, we conducted frequently sampled intravenous glucose tolerance tests (FSIGT) to assess pharmacodynamic responses to a rapid-acting GLP1R agonist (exenatide): glucose-stimulated insulin secretion and the rate of glucose disappearance. We applied the “minimal model” to analyze FSIGT data. This mathematical model describes the time course for plasma glucose levels to return to baseline after an intravenous glucose challenge as a function of the time course of glucose-stimulated insulin secretion and two parameters: insulin sensitivity (S_i_) and glucose effectiveness (S_g_) ^8^. S_g_ reflects insulin-independent mechanisms whereby plasma glucose concentration drives the return of plasma glucose to baseline levels during an FSIGT. Herein we summarize pharmacodynamic responses to exenatide in the overall study population as well as in sub-groups who were homozygous for two genetic variants, *GIPR* (p.E354Q; rs1800437) and *GCGR* (p.G40S; rs850763). The choice of these two candidate genes was inspired by intriguing observations suggesting interactions among GLP1, GIP, and glucagon ^9^. Specifically, peptide drugs that target multiple receptors (GLP1R, GIPR, and GCGR) are reported to exert stronger pharmacological effects than selective drugs targeting only one receptor ^9^. Just as exogenous administration of agonists targeting GIPR and GCGR enhance pharmacological responses to an exogenous GLP1R agonist, we hypothesized that response to an exogenous GLP1R agonist might be modulated by effects of endogenous agonists mediated by GIPR and/or GCGR. We focused on two variants (rs1800437 and rs850763), which have been reported to alter the function of GIPR and GCGR, respectively ^10, 11^.

## 2 | Methods

### 2.1 | Study population: recruitment and screening

The Old Order Amish population of Lancaster County, PA emigrated from Central Europe in the early 1700’s. University of Maryland School of Medicine researchers have been studying genetic determinants of cardiometabolic health in this population since 1993. To date, ∼10,000 Amish adults participated in one or more studies as part of the Amish Complex Disease Research Program (http://www.medschool.umaryland.edu/endocrinology/Amish-Research-Program/). These studies generated a genotype database that was used to identify individuals with any of three genotypes who were eligible for our clinical trial: (a) homozygotes for a missense variant in *GIPR* (p.E354Q; rs1800437); (b) homozygotes for a missense variant in *GCGR* (p.G40S; rs850763); and (c) individuals who were homozygous for the “wild type” major alleles of *GIPR* and *GCGR*. A research nurse, accompanied by a member of the Amish community, made home visits to invite individuals to participate in the study. If they expressed interest, the study was explained in detail; potential participants were invited to sign an informed consent form. Thereafter, the research nurse obtained a medical history; measured height, weight and blood pressure; and obtained blood samples for screening laboratory tests (hematocrit, fasting plasma glucose, serum creatinine, serum AST, serum ALT, plasma TSH, and HbA1c).

### 2.2 | Study design: overview

The study was designed as a crossover study in which healthy individuals were randomized between administration of exenatide (5 µg, sc) or saline (0.2 mL, sc). Fifteen minutes after the injections, participants underwent a frequently sampled intravenous glucose tolerance test (FSIGT) as described in the Supplementary Appendix. A second FSIGT was conducted 5-28 days later; each participant received the treatment (either saline or exenatide) not administered for the first FSIGT. All procedures were reviewed and approved by the University of Maryland Baltimore IRB.

Practical considerations guided our selection of exenatide as the GLP1R agonist for this pilot study. Exenatide is rapidly absorbed and could be administered 15 min prior to initiation of the FSIGT. In contrast, liraglutide (the principal alternative in 2015 when this study was initiated) achieves Cmax ∼10-12 hours after injection ^12^. The logistics of administering an injection 10-12 hours prior to the FSIGT would have been challenging in this out-patient study.

### 2.3 | Power calculations

We aimed to recruit 24 research participants for each of the three genotypes. We estimated that these recruitment targets provided 80% power (with p=0.05) to detect association with a 35% change in first-phase insulin secretion and a 25% change in glucose disappearance rate for homozygous variant compared to homozygous wild-type individuals. However, the clinical trial was terminated early when it became apparent that we would not meet our recruitment targets within the available budget.

### 2.4 | Eligibility criteria

To be eligible to participate in the clinical trial, individuals were required to be of Amish descent, at least 18 years old, and have BMI between 18-40 kg/m^2^. Exclusion criteria are summarized in the Supplementary Appendix.

### 2.5 | Clinical chemistry

Processing of blood samples and laboratory methods are summarized in the Supplementary Appendix.

### 2.6 | Data and statistical analyses

We established two primary end points: drug effects on (a) first phase insulin secretion and (b) glucose disappearance rate. Fig. 1 presents untransformed data on plasma levels of insulin and glucose. Tables 1 and 2 present analyses of logarithmic transformation of data on AIR_g_, S_i_, S_g_, and glucose disappearance rates because the untransformed data were not normally distributed. Data in all the other Figures and Tables were inverse normally transformed and multiplied by SD to have effect size in real units; all analyses in those Figures and Tables were adjusted for age, sex, and BMI. Genetic association analysis (Table S3) was performed using linear mixed models to account for familial correlation using the kinship relationship matrix as implemented in the Mixed Model Analysis for Pedigree and population [https://mmap.github.io/]. Multiple regression analysis was conducted using Excel. Minimal model analyses were conducted as described elsewhere ^13–15^. Minimal model analysis of the FSIGT data was used to estimate indices of glucose effectiveness (S_g_) and insulin sensitivity (S_i_) as previously described using MINMOD software (version 6.02) (MinMOD Millenium, Los Angeles, CA) ^15^. AIR_g_ is calculated as the area under the curve for first phase insulin secretion between 0-10 minutes. Disposition Index was calculated as the product of S_i_ multiplied by AIR_g_ ^8^.

**Figure 1.**
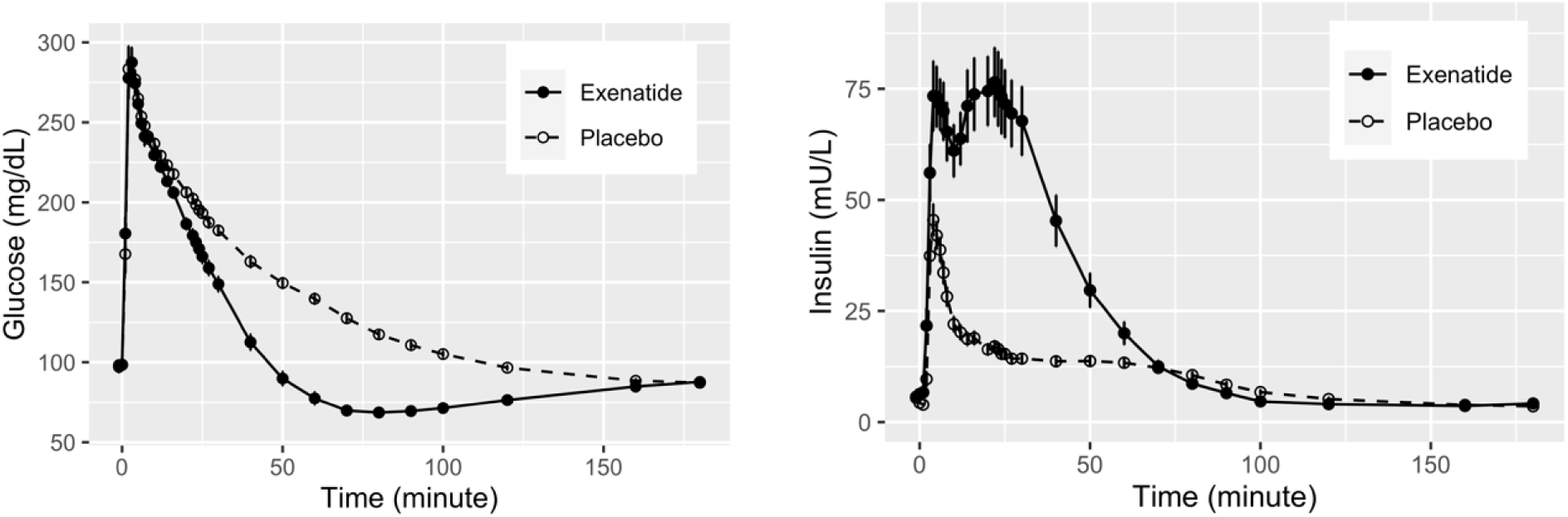
Exenatide enhances insulin secretion and accelerates the rate of glucose disappearance decrease in frequently sampled intravenous glucose tolerance tests (FSIGT). Fifty-three non-diabetic healthy volunteers participated in a randomized crossover study of exenatide (N=53). After an overnight fast, participants were randomly assigned to receive either exenatide (5 µg, s.c.; solid circles) or saline (0.2 mL, s.c.; open circles). The FSIGT was initiated 15 min after the s.c. injection by administration of glucose (0.3 g/kg, i.v.) over two minutes. Blood samples were obtained for measurement of plasma glucose (panel A) and plasma insulin (panel B) over the course of 180 min after glucose administration as described in the Methods section ^8, 13, 14, 17^. After a washout period of 5-28 days, a second FSIGT was conducted in which the participant received the treatment (either saline or exenatide) not administered for the first FSIGT. Data are presented as means ± SEM.

**Table 1.** Parameters from minimal model analyses. This Table summarizes data from 45 frequently sampled intravenous glucose tolerance tests conducted after research participants received either saline (placebo) or exenatide. Data are presented as means ± SEM. The first five rows present analyses of untransformed data whereas the next five rows present analyses of logarithmically transformed data. Paired t-tests were conducted on the logarithmically transformed data; two-sided p-values are presented in the last column of the Table. Rates of glucose disappearance were calculated as described in Table S2. Except for one participant with Si of 0.16, the remaining participants’ S_i_ values ranged from 1.4-17.2 × 10^−4^ ⋅ min^−1^ /[µU/mL] as compared to a published reference range of 1.4-25.7 × 10^−4^ ⋅ min^−1^ / [µU/mL] in lean individuals ^18^.

| Parameter | Placebo | Exenatide | p-value |
| --- | --- | --- | --- |
| **Insulin sensitivity ($S_i$)<sup>a</sup>** | $6.80 \pm 0.60$ | $7.07 \pm 0.58$ | |
| **Glucose effectiveness ($S_g$)<sup>b</sup>** | $0.0139 \pm 0.0010$ | $0.0183 \pm 0.010$ | |
| **Acute insulin response ($\text{AIR}_g$)<sup>c</sup>** | $243 \pm 24$ | $472 \pm 56$ | |
| **Disposition index (DI)** | $1454 \pm 157$ | $2833 \pm 288$ | |
| **Glucose disappearance rate<sup>d</sup>** | $506 \pm 31$ | $1060 \pm 70$ | |
| **Log ($S_i$)** | $0.73 \pm 0.05$ | $0.78 \pm 0.04$ | 0.36 |
| **Log ($S_g$)** | $-1.92 \pm 0.04$ | $-1.76 \pm 0.03$ | 0.0008 |
| **Log ($\text{AIR}_g$)** | $2.31 \pm 0.04$ | $2.58 \pm 0.04$ | $2.3 \times 10^{-10}$ |
| **Log (DI)** | $3.04 \pm 0.05$ | $3.36 \pm 0.04$ | $2.0 \times 10^{-6}$ |
| **Log (glucose disappearance rate)** | $2.66 \pm 0.03$ | $2.96 \pm 0.04$ | $1.4 \times 10^{-13}$ |

<sup>a</sup> units: $10^{-4} \cdot \text{min}^{-1} / [\mu\text{U/mL}]$
<sup>b</sup> units: $\text{min}^{-1}$
<sup>c</sup> units: $\mu\text{U} \cdot \text{mL}^{-1} \cdot \text{min}^{-1}$
<sup>d</sup> units: $10^{-5} \cdot \text{min}^{-1}$

**Table 2.**
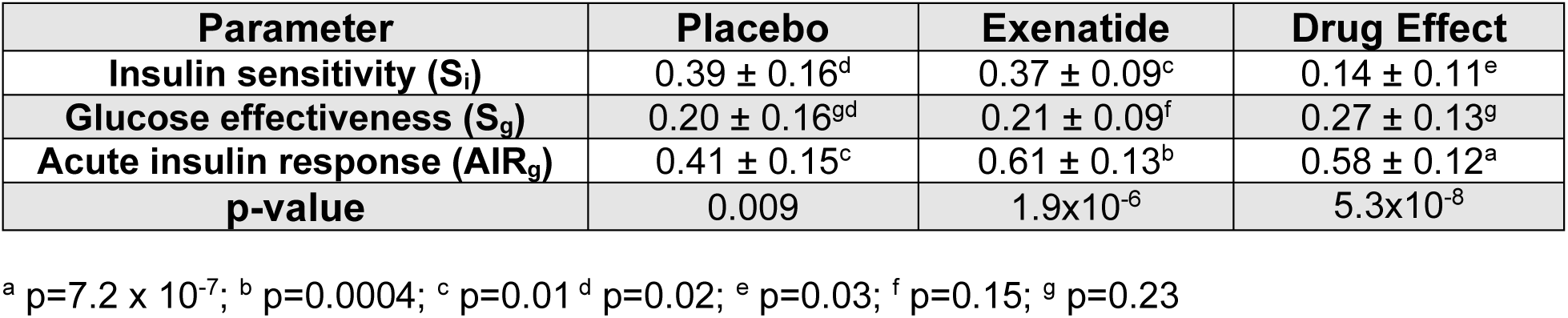
Relative contributions to the rate of glucose disappearance: beta coefficients from multiple regression of parameters derived from minimal model analysis. The Table presents results of multiple regression analyses with three independent variables derived from minimal model analysis of frequently sampled intravenous glucose tolerance tests. The first two columns present data on logarithmically transformed values of insulin sensitivity (S_i_), glucose effectiveness (S_g_), and acute insulin response to glucose (AIR_g_) in studies where participants received either saline (placebo) or exenatide. The last column presents analysis of logarithmically transformed data on effects of exenatide (i.e., “drug effects” calculated as specified in Table S2) on insulin sensitivity (S_i_), glucose effectiveness (S_g_), and acute insulin response to glucose (AIR_g_). in each analysis, the dependent variable was the rate of glucose disappearance as defined in Table S2. Beta values are expressed in standard deviation units (SDU) – i.e., the change in the dependent variable relative to the change in the independent variable in SDU. The last row of the Table presents the overall two-sided p-value for each multiple regression model as calculated using Excel. The footnotes indicate two-sided p-values for each of the coefficients. Units for the minimal model parameters are summarized in Table 1.

Statistical significance was assessed using a paired t-test. Although a p-value of p<0.05 was defined as the threshold for nominal statistical significance, a more stringent threshold (e.g., p<0.001) may be appropriate to account for multiple comparisons.

## 3 | Results

### 3.1 | Disposition and adverse events

Research participants’ baseline characteristics are summarized in Table S1. Disposition of participants and adverse events are summarized in Fig. S1. Seventy-eight individuals were enrolled in this clinical trial between June, 2016 – November, 2018. Sixty-two participants completed frequently sampled intravenous glucose tolerance tests.

### 3.2 | Intravenous glucose tolerance tests: impact of exenatide

Intravenous administration of glucose triggered prompt increases in plasma levels of glucose and insulin (Fig. 1). Plasma glucose achieved a peak at ∼4 min – after which time glucose levels decreased. Administration of exenatide augmented insulin secretion. We defined indices to quantitate exenatide’s direct effect on glucose-stimulated insulin secretion and exenatide’s indirect effect on the rate of glucose disappearance (Table S2). As illustrated by the time course of insulin levels in exenatide-treated individuals (Fig. 1), insulin secretion was biphasic. Consistent with published literature ^12, 16^, the area-under-the-curve (AUC) between 0-10 min provides an index of first phase insulin secretion (Table S2). We used AUC between 10-50 min as an index of second phase insulin secretion. The higher plasma insulin levels observed in exenatide-treated individuals accelerated the rate at which glucose levels declined. During the time interval between 25-50 min, the plot of log(glucose) versus time approximates a straight line. The slope of that line served as index of the rate of glucose disappearance (Table S2) ^12^. Exenatide triggered statistically significant increases in the areas under the curve (AUCs) for first phase (∼1.9-fold; p=1.9×10^-9^; Fig. S2) and second phase (∼3.5-fold; p=1×10^-9^) insulin secretion as well as the rate of glucose disappearance (∼2.4-fold; p=2×10^-10^; Fig. S2). Values for first phase and second phase insulin secretion were closely correlated (r=0.74, p=2.4×10^-10^ for placebo data; r=0.63, p=3.4×10^-7^ for data after administration of exenatide) (Fig. S3).

Because curves describing time courses for mean glucose levels in the placebo and exenatide studies are essentially superimposable during the first ten minutes (Fig. 1), the drug effect on first phase insulin secretion likely represents a direct effect of exenatide. In contrast, exenatide’s effect on second phase insulin secretion (10-50 min) likely reflects a balance between exenatide’s positive direct effect on insulin secretion and the negative impact of lower glucose levels tending to decrease insulin secretion. Accordingly, we emphasized first phase insulin secretion to assess the correlation between insulin secretion and the rate of glucose disappearance. The rate of glucose disappearance (as defined in Table S2) was correlated with first phase insulin secretion (r=0.74, p=2.4 × 10^-10^ for placebo data; r=0.63, 3.4×10^-7^ for data after administration of exenatide) (Fig. 2AB). As illustrated in the histograms (Fig. S4), we observed substantial inter-individual variation in indices for drug effects on both stimulation of insulin secretion and acceleration of glucose disappearance. While exenatide exerted little or no effect in some individuals, greater than 5-fold increases were observed in other individuals.

### 3.2 | Intravenous glucose tolerance tests: exenatide increases glucose effectiveness

Exenatide’s effect on the rate of glucose disappearance was correlated with the magnitude of exenatide’s effect on insulin secretion (r=0.70; p=9.2×10^-9^) (Fig. 2C). The correlation coefficient suggests that inter-individual variation in the magnitude of exenatide’s direct effect on insulin secretion explains approximately half of the observed variance in the magnitude of exenatide’s indirect effect on the rate of glucose disappearance. Accordingly, we searched for other factors that might have contributed to variation in the effect of exenatide to accelerate glucose disappearance. We applied Bergman’s minimal model ^8, 15, 17, 18^ to estimate insulin sensitivity (S_i_), glucose effectiveness (S_g_), and disposition index (DI) (Table 1). Although acute administration of exenatide did not alter insulin sensitivity (p=0.36), exenatide treatment increased glucose effectiveness by 32% (p=0.0008), acute insulin response to glucose by 94% (p=2.3×10^-10^) and disposition index by 95% (p=2.0×10^-6^). The rate of glucose disappearance was significantly correlated with glucose effectiveness for data from both placebo studies (r=0.50, p=0.0001; Fig. 3) and exenatide studies (r=0.66, p=5.4×10^-8^; Fig. 3). We conducted multiple regression analyses to further investigate inter-relationships among the parameters of the minimal model in the context of acute administration of exenatide. In the placebo state, inter-individual variation in in S_i_ or AIR_g_ each made similar magnitude contributions to inter-individual variation the rate of glucose disappearance. An increase of 1 standard deviation unit (SDU) in S_i_ or AIR_g_ was associated with an increase of ∼0.4 SDU in the rate of glucose disappearance (Table 2). Inter-individual variation in glucose effectiveness made a smaller contribution (i.e., 0.2 SDU increase in glucose disappearance rates for each SDU increase in S_g_) (Table 2). After exenatide administration, variation in all three parameters also contributed, but the effect of variation in AIR_g_ predominated (an increase of 0.61 SDU in the rate of glucose disappearance per 1 SDU increase in insulin secretion); the impact of variations in insulin sensitivity and glucose effectiveness were smaller (Table 2). Variation in the magnitude of the drug effect on insulin secretion was the principal determinant of the magnitude of the drug effect on the rate of glucose disappearance. An increase of 1 SDU in the drug effect on AIR_g_ was associated with an increase of 0.58 ± 0.12 SDU in the magnitude of the drug effect for the rate of glucose disappearance (p=7.2 × 10^-7^) (Table 2).

**Figure 2.**
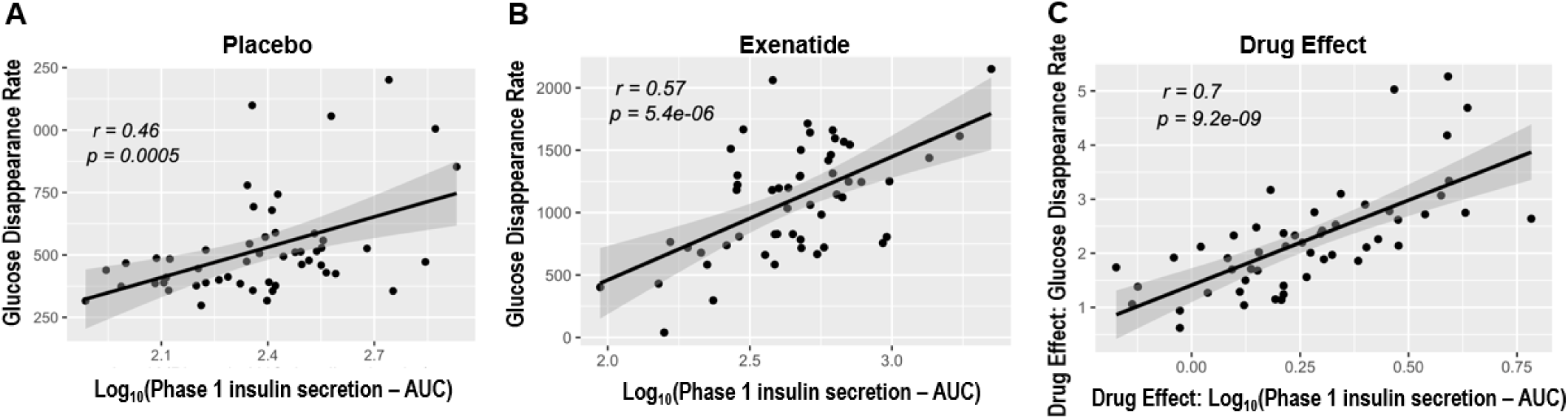
Correlations between the rate of glucose disappearance and first phase insulin secretion. Glucose-stimulated insulin secretion accelerates the rate of glucose disappearance, which in turn provides an “off” signal for insulin secretion. This analysis demonstrates that the index for the rate of glucose disappearance between 25-50 min (Table 1) is correlated with the index for first phase insulin secretion (Table 1) in both the placebo-FSIGT (panel A; r=0.74; p=2.4×10^-10^) and the exenatide-FSIGT (panel B; r=0.63; p=3.4×10^-7^). Indices for effects of exenatide on first phase insulin secretion and the rate of glucose disappearance are defined in Table 1. The index for exenatide-induced acceleration of glucose disappearance is correlated with the index for exenatide-induced insulin secretion (panel C; r=0.70; p=9.2×10^-9^). Indices for 1^st^ phase insulin secretion are expressed as logarithms of AUC expressed in the following units: [(µU/mL) · min]. Indices for glucose disappearance rates are expressed in the following units: 10^-5^ · min^-1^].

**Figure 3.**
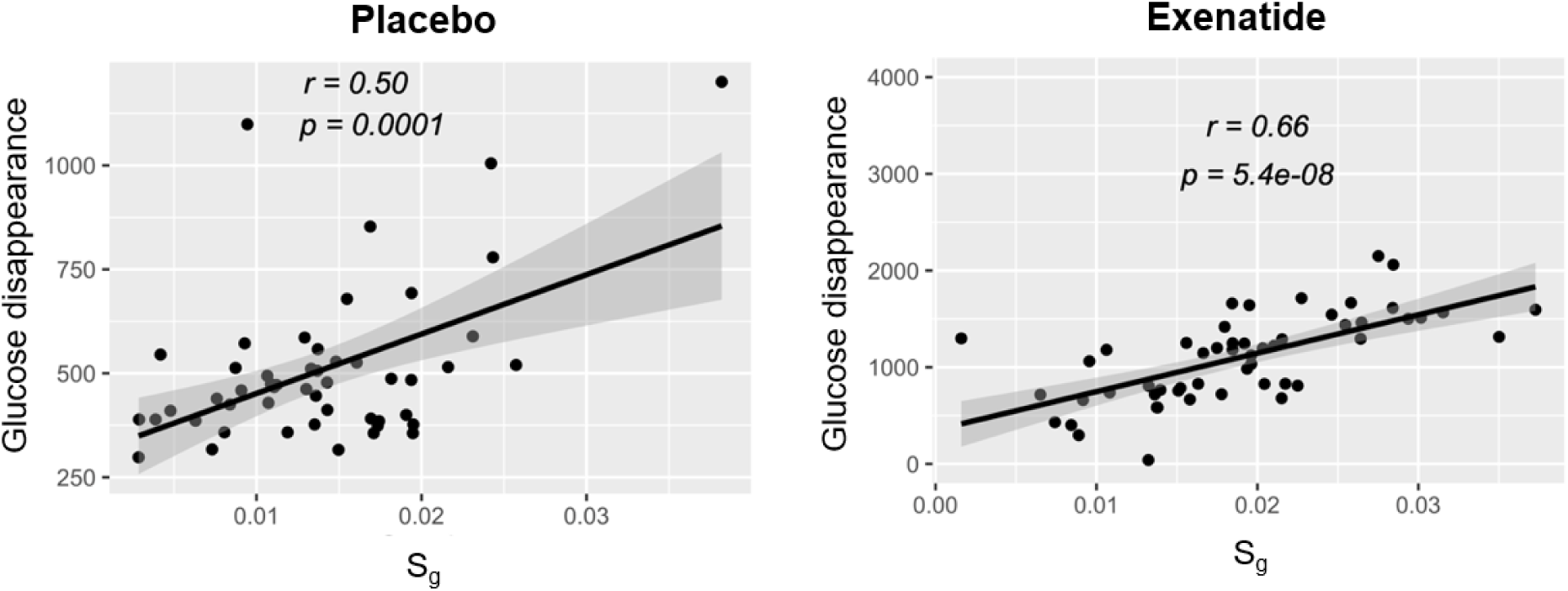
Correlation of rate of glucose disappearance with glucose effectiveness (Sg). Rates of glucose disappearance (vertical axis) are plotted as a function of glucose effectiveness (S_g_) (horizontal axis). The correlation coefficients and p-values are indicated in the Figures for the placebo studies (left panel) and the exenatide studies (right panel). Values of S_g_ are expressed in the following units: min^-1^. Rates of glucose disappearance were calculated as summarized in Table S2 expressed in the following units (10^-5^ · min^-1^).

### 3.3 | Effect of genotype

This pilot study was designed to test the hypothesis that missense variants in two candidate genes (*GCGR* and *GIPR*) might be associated with pharmacodynamic responses to GLP1 receptor agonists. Table S3 compares pharmacodynamic responses to exenatide among three groups of people: (a) individuals who are homozygous for the major alleles of both genes; (b) homozygotes for the G40S variant of *GCGR* (rs1801483) ^11, 19^; and (c) homozygotes for the E354Q variant of *GIPR* (rs1800437) ^10, 20, 21^ (Table S3). Four co-primary endpoints were pre-specified: association of genotype (either G40S-*GCGR* or E354Q-*GIPR*) with the magnitude of drug effect (i.e., either exenatide-induced augmentation of first phase insulin secretion or exenatide-induced acceleration of the rate of glucose disappearance). We did not observe statistically significant associations between any of the genetic variants and either of the endpoints.

## 4 | Discussion

Pharmacogenomic research has utilized varied approaches – including acute studies in healthy volunteers assessing acute pharmacodynamic endpoints ^3, 22^ and chronic studies in disease patients assessing routine clinical endpoints such as HbA1c ^23^. Short-term studies in healthy volunteers offer several methodological advantages – for example, minimization of confounding factors caused by co-existing diseases or effects of changing co-medications during the course of the study. This pilot study provided an opportunity to validate pharmacodynamic endpoints for our currently ongoing genome-wide association study of the genetics of pharmacodynamic responses to a GLP1 receptor agonist (NCT05071898). As exemplified by the usual stepwise approach to drug development, it is informative to conduct pilot studies to validate methods and analytic approaches before initiating a larger clinical trial. Our results confirm the existence of substantial inter-individual variation in pharmacodynamic responses to exenatide – ranging from no significant increase to a 5-to 6-fold increase for glucose-sensitive insulin secretion or acceleration of glucose disappearance, respectively. The wide range of inter-individual variation in the magnitude of drug response emphasizes the potential value of identifying genetic variants to predict an individual’s pharmacological response and provide guidance to physicians in selecting the best diabetes drug for each individual patient.

FDA-approved prescribing information for GLP1R agonists identifies several important clinical benefits for this class of drugs: HbA1c-lowering in patients with type 2 diabetes, weight loss in overweight/obese patients, and decreasing major adverse cardiovascular events in patients with type 2 diabetes ^24–26^. Our clinical trial assessed drug-induced acceleration of glucose disappearance in healthy non-diabetic individuals, which is mechanistically related to drug-induced HbA1c lowering in patients with type 2 diabetes. Mediation analysis of the LEADER cardiovascular outcome trial suggested that ∼90% of liraglutide’s beneficial impact on cardiovascular risk reduction was mediated by the drug-induced decrease in HbA1c ^27^. This suggests that genetic variants associated with glycemic effects may also contribute to predicting the magnitude of cardiovascular risk reduction. Comparative effectiveness confirmed that GLP1R agonists providing the largest HbA1c-lowering also provided the greatest cardioprotection ^28, 29^.

### 4.1 | Acute exenatide-induced acceleration of glucose disappearance: mechanisms

Exenatide increased the magnitude of the mean glucose-stimulated 1^st^ phase insulin secretion by 1.9-fold, 3.5-fold increase for 2^nd^ phase insulin secretion, and 2.1-fold increase for the glucose disappearance rate (Figs. 1 and S2; Table 1). Furthermore, exenatide caused a 32% increase in mean glucose effectiveness (S_g_) but did not induce a significant change in mean insulin sensitivity (Table 1). The impact of the exenatide-induced increase in S_g_ is supported by the observation that rates of glucose disappearance were correlated with the magnitude of S_g_ in both placebo studies and studies conducted after participants received exenatide (Fig. 3).

Multiple regression analysis revealed that inter-individual variation in the magnitudes of glucose-stimulated insulin secretion and insulin sensitivity appeared to contribute significantly to inter-individual variation in the rate of glucose disappearance in both placebo studies and studies conducted after participants received exenatide (Table 2). Thus, although exenatide does not exert an acute effect on insulin sensitivity, pre-existing inter-individual variation in S_i_ does contribute significantly to inter-individual variation in rates of glucose disappearance both at baseline and in the exenatide-treated states (Table 2). Notwithstanding smaller contributions from S_g_ and S_i_, inter-individual variation in the magnitude of exenatide-induced increase in glucose-sensitive insulin secretion (AIR_g_) was quantitatively the most important determinant of the magnitude of the drug-induced increase in glucose disappearance (Table 2); a change of 1 SDU in the drug effect on AIR_g_ was associated with a 0.58 SDU in the drug effect on the rate of glucose disappearance. These data on effects of exenatide on sub-phenotypes (i.e., insulin secretion and glucose disappearance rate) are generally similar to what has been reported for liraglutide and lixisenatide ^12, 16^ with the exception that lixisenatide was reported not to induce a statistically significant increase in 2^nd^ phase insulin secretion in healthy individuals ^16^. Consistent with previous studies of GLP1R agonists ^12, 16, 30, 31^, we did not detect a significant effect of exenatide on the mean insulin sensitivity index (S_i_). The selectivity of the exenatide-induced increase in S_g_ without affecting S_i_ supports the conclusion that S_i_ and S_g_ reflect distinct aspects of glucose metabolism ^8^.

The term “glucose effectiveness” refers to insulin-independent mechanisms whereby glucose concentrations enhance glucose utilization and/or decrease glucose production ^8^. The precise physiological mechanisms mediating glucose effectiveness are not completely understood ^8^. There has been discussion about challenges in estimating glucose effectiveness from data derived from intravenous glucose tolerance tests. For example, it has been suggested that higher levels of insulin secretion may lead to higher mathematical estimates of glucose effectiveness; it has been challenging to identify detailed mechanisms accounting for these higher estimates of S_g_ ^32, 33^. Nevertheless, it is theoretically possible that the exenatide-induced increase in insulin secretion may contribute to the larger mathematical estimates of S_g_. Bergman ^8^ has suggested that glucose effectiveness may reflect hepatic glucose uptake and metabolism (mediated by GLUT2 and glucokinase). As GLP1 receptor agonists have been reported to suppress glucagon levels ^34^, this might provide an endocrine mechanism whereby exenatide might exert an indirect effect on hepatic glucose metabolism. However, to the extent that effects of exenatide on glucagon secretion by α-cells might be mediated at least in part by paracrine β-cell insulin secretion, such an effect would not necessarily be entirely independent of insulin. In any case, further research will be required to elucidate physiological mechanisms mediating acute effects of GLP1 receptor agonists on glucose effectiveness.

### 4.2 | Pharmacogenomic candidate genes

A missense variant in the *GLP1R* gene (rs6923761; p.T149M) has previously been reported to be associated with a diminished response to GLP1 receptor agonists ^6, 7, 35^. We have not detected this variant in DNA sequences in the Old Order Amish. Recent literature has suggested the existence of positive crosstalk among GLP1, GIP, and glucagon with respect to their beneficial effects on insulin secretion and weight loss ^36^. These considerations led to the design of tirzepatide – a recently approved “twincretin” combining agonist activity against both GLP1R and GIPR ^37^. Based on these data, we hypothesized that there might be pharmacologically relevant interactions between an exogenous GLP1R agonist and endogenous glucagon or GIP. To test these hypotheses, we recruited individuals who are homozygous for two relatively common missense variants: rs1801483 (p.G40S) in *GCGR* and rs1800437 (p.E354Q) in *GIPR*. The p.E354Q variant in *GIPR* has been reported to enhance agonist-induced receptor desensitization – potentially leading to resistance to the effect of GIP agonists ^10^. Furthermore, this genetic variant in *GIPR* has been reported to be associated with increased plasma levels of GIP as well as various anthropometric and glycemic traits ^38^. The p.G40S variant in *GCGR* has been reported to impair signaling through β-arrestin-1 ^11^. This p.G40S variant in *GCGR* has been reported to be associated with diabetes ^19^ although this finding has not been reproducible ^39^.

Although we observed 15-20% numerical decreases in the magnitudes of the effect of exenatide-induced effect on second phase glucose-stimulated insulin secretion in homozygotes for variants in both *GIPR* (rs1800437) and *GCGR* (rs1801483), these numerical differences were not statistically significant (Table S3). We encountered greater than expected difficulty in recruiting research participants with genotypes of interest. The small size of the study population creates uncertainty about the interpretation of our observations – i.e., whether the statistically insignificant numerical trends toward a decrease in the response to exenatide might represent type II error (incorrect rejection of a false null hypothesis). In order to increase power to detect genetic associations, this study focused on homozygotes. Heterozygosity is quite a bit more common and would, therefore, be of greater practical relevance from a pharmacogenomic perspective. Even if one assumes that these variants in GIPR or GCGR are indeed associated with responses to GLP1R agonists, we conclude that heterozygosity for these variants is unlikely to exert a large, clinically important effect on responses to GLP1R agonists. Further research will be required to resolve these uncertainties. Whether or not the missense variant in *GIPR* alters the response to a selective GLP1R agonist, it would be of considerable interest to determine whether the variant in *GIPR* is associated with variation in response to peptides possessing GIPR agonist activity (including tirzepatide) ^37^.

## Data Availability

Data is available to qualified academic researchers subject to a data transfer agreement to protect participants' confidential information.

## ACKNOWLEDGEMENTS

We gratefully acknowledge funding provided by the American Diabetes Association (grant number 1-16-ICTS-112) and National Institute of Diabetes and Digestive and Kidney Disease (R01DK130238, T32DK098107, and P30DK072488). We also gratefully acknowledge contributions of the research participants and the skilled staff at the Amish Research Clinic for their critical roles in making this study possible. We are grateful to Mary Pavlovich, Melanie Daue, and Kathy Ryan for their help with database management. Dr. Laura Yerges-Armstrong provided genotype data enabling us to identify individuals to be invited to participate in this genotype-guided recruitment study.

## AUTHORS’ CONTRIBUTIONS

*<u>Conception of the clinical trial and PI for the grant from the American Diabetes Association grant</u> (*1-16-ICTS-112): SIT

*<u>Acquisition and analysis of data</u>*: ALB, AHY, MEM, EAS, SIT, HBW, ZSY

*<u>Minimal model analysis of FSIGT data</u>*: RM, HF

*<u>Establishment of Old Order Amish genotype database</u>*: BDM, ARS

*<u>Preparation of first draft of manuscript</u>*: SIT

*<u>Revising and approving final version of manuscript</u>*: all authors

*<u>Overall accountability for all aspects of work</u>*: SIT

## Supplementary Appendix

### 1 | Exclusion Criteria

- Known allergy to exenatide
- History of diabetes, random glucose >200 mg/dL, or HbA1c ≥ 6.5%
- Significant cardiac, hepatic, pulmonary, or renal disease or other diseases that the investigator judged would make interpretation of the results difficult or increase the risk of participation
- Seizure disorder
- Pregnant by self-report or known pregnancy within 3 months of the start of study
- Currently breast feeding or breast feeding within 3 months of the start of the study
- Estimated glomerular filtration rate <60 mL/min/1.73m^2^
- Hematocrit <35%
- Liver function tests greater than 2 times the upper limit of normal
- Abnormal TSH
- History of pancreatitis or pancreatic cancer. Personal or family history of medullary carcinoma of the thyroid.

### 2 | Frequently sampled intravenous glucose tolerance tests

After an overnight fast, participants were transported to the Amish Research Clinic where FSIGTs were performed (1Fo, 2). Two 20-gauge catheters were placed in antecubital veins through which normal saline (60 mL/hr) was administered – one for intravenous administration of D50W and one for obtaining blood samples. The infusion was stopped at times when blood samples were withdrawn. To minimize dilution of blood, small amounts of blood were discarded prior to obtaining blood samples. After placement of intravenous catheters, participants received subcutaneous injections of either exenatide (5 µg, sc) or saline (0.2 mL, sc). Fifteen minutes after the subcutaneous injections, an intravenous bolus of glucose (0.3 g/kg) was infused over two minutes. Thirty-one timed blood samples were obtained between –10– and +180-minutes relative to the i.v. glucose infusion (1). Plasma samples were used to assay levels of glucose and insulin.

### 3 | Clinical Chemistry

Screening blood samples were obtained by a research nurse during home visits and collected in test tubes as appropriate for each assay: EDTA anticoagulant (purple top tube) for measurement of hematocrit and HbA1c; heparin anticoagulant (green top tube) for measurement of TSH; gray top tubes containing sodium fluoride and potassium oxalate for measurement of fasting plasma glucose; red top tube for collecting serum samples. After placing gray, purple, and green top tubes on ice, blood samples were transported to the clinical laboratory at the Amish Research Clinic (maximum transport time, 2 hours). After centrifugation (3300 rpm for 10 min), plasma/serum was sent on the same day to Quest Diagnostics for assay. Blood samples for the frequently sampled intravenous glucose tolerance test were collected in EDTA-containing purple top tubes for measurement of plasma insulin and in EDTA/oxalate-containing gray top tubes for measurement of plasma glucose. Glucose was measured in duplicate using an YSI glucose analyzer. Insulin was assayed in duplicate following the manufacturer’s directions using reagents in kits (#10-1113-01) purchased from Mercodia Inc.

**Table S1.** Demographics and baseline characteristics of study population.

| **Mean ± SEM (range)** | **Males** | **Females** | **Total** |
| --- | --- | --- | --- |
| **N (sample size)** | 33 | 21 | 54 |
| **Age (years)** | 46.0 ± 2.0 | 52.0 ± 2.1 | 48.3 ± 1.5 |
| **BMI (kg/m<sup>2</sup>)** | 27.3 ± 0.6 | 28.9 ± 0.8 | 27.9 ± 0.5 |
| **HbA1c (%)** | 5.51 ± 0.6 | 5.60 ± 0.7 | 5.54 ± 0.4 |
| **Serum creatinine (mg/dL)** | 0.88 ± 0.02 | 0.73 ± 0.02 | 0.82 ± 0.02 |
| **eGFR (mL/min/1.73 m<sup>2</sup>)** | 101.2 ± 2.2 | 94.5 ± 3.4 | 98.6 ± 1.9 |
| **Hematocrit (%)** | 43.6 ± 0.4 | 39.0 ± 0.5 | 41.8 ± 0.4 |
| **Aspartate amino transferase (U/L)** | 18.5 ± 0.8 | 17.4 ± 0.9 | 18.1 ± 0.6 |
| **Alanine amino transferase (U/L)** | 21.0 ± 1.4 | 17.3 ± 1.1 | 19.6 ± 1.0 |

**Table S2.** Definition of parameters for statistical analyses.

| Parameter | Definition |
| --- | --- |
| 1<sup>st</sup> phase insulin secretion | Area under the curve (AUC) for plasma insulin levels (0-10 min) calculated by trapezoidal rule |
| 2<sup>nd</sup> phase insulin secretion | Area under the curve for plasma insulin levels (10-50 min) calculated by trapezoidal rule |
| Glucose disappearance rate | Slope of the plot of the logarithm of glucose concentration versus time (25-50 min) calculated as the slope of the least square fit using Excel. |
| Drug effect: 1<sup>st</sup> phase insulin secretion | Logarithm of AUC for 1<sup>st</sup> phase insulin secretion when participant received exenatide – logarithm of AUC for 1<sup>st</sup> phase insulin secretion when participant received saline. |
| Drug effect: 2<sup>nd</sup> phase insulin secretion | Logarithm of AUC for 2<sup>nd</sup> phase insulin secretion when participant received exenatide *minus* logarithm of AUC for 2<sup>nd</sup> phase insulin secretion when participant received saline. |
| Drug effect: glucose disappearance rate | Glucose disappearance rate when participant received exenatide *minus* the glucose disappearance rate when participant received saline. |
| Log-transformed glucose disappearance rate | Logarithm of slope of the plot of the logarithm of glucose concentration versus time (25-50 min) calculated as the slope of the least square fit using Excel. |
| Drug effect: log-transformed glucose disappearance rate | Logarithm of glucose disappearance rate when participant received exenatide *minus* logarithm of the glucose disappearance rate when participant received saline. |

**Table S3.**
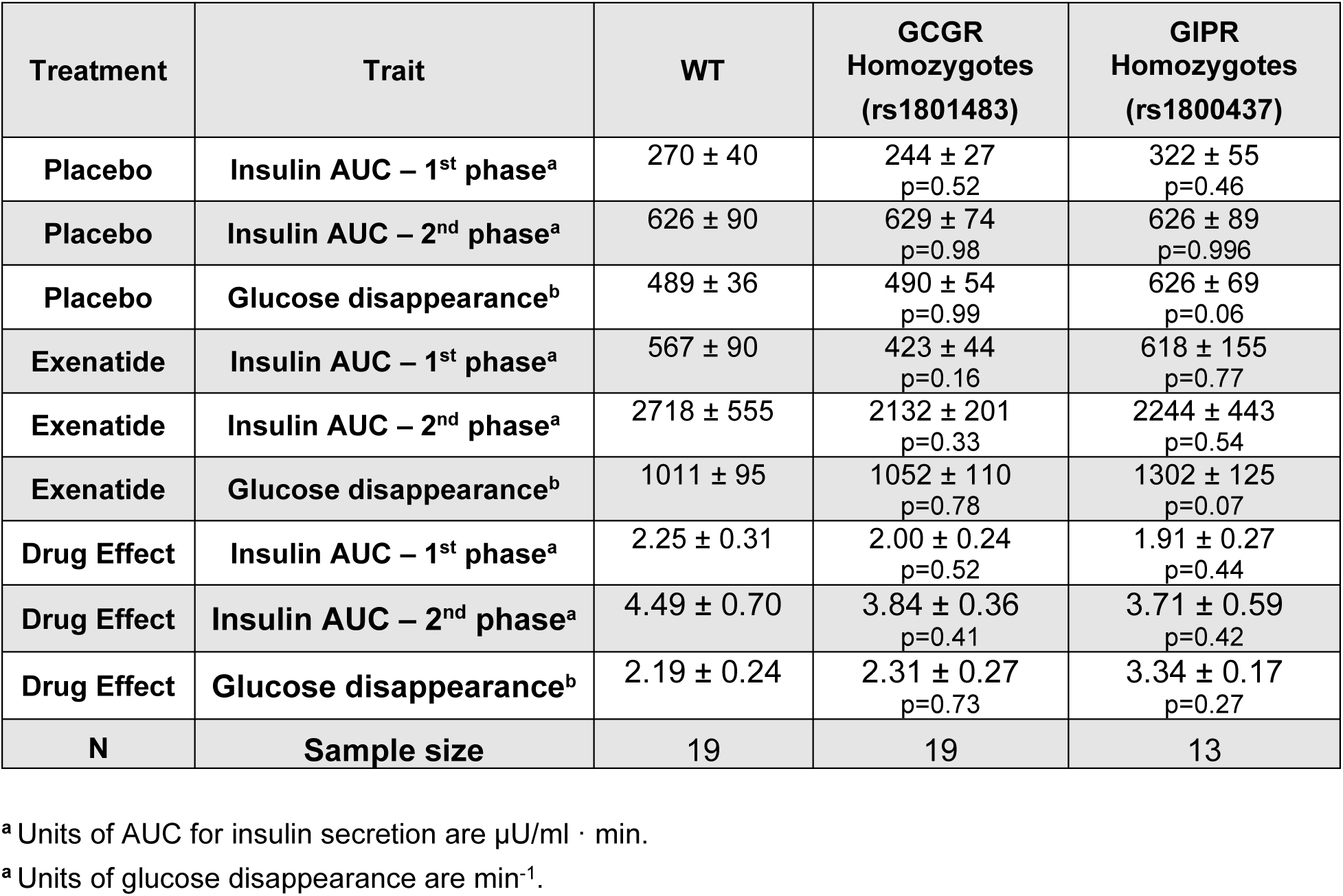
Interactions of genotype with pharmacodynamic responses to exenatide. Data are expressed as means ± SEM. Insulin secretion was calculated as areas under the curve (AUC) using the trapezoidal rule for times between 0-10 min (1^st^ phase) and 10-50 min (2^nd^ phase). Three participants were excluded because their genotypes did not correspond to any of the three groups. Two participants were homozygous for rs1801483 and heterozygous for rs1800437; one participant was homozygous for rs1800437 and heterozygous for rs1801483. One participant who was homozygous for rs1801483 was excluded because of missing data. P-values were calculated using Student’s t-test for paired values. To account for the 18 statistical comparisons in the Table, p<0.0025 was chosen as the criterion for statistical significance.

**Figure S1.**
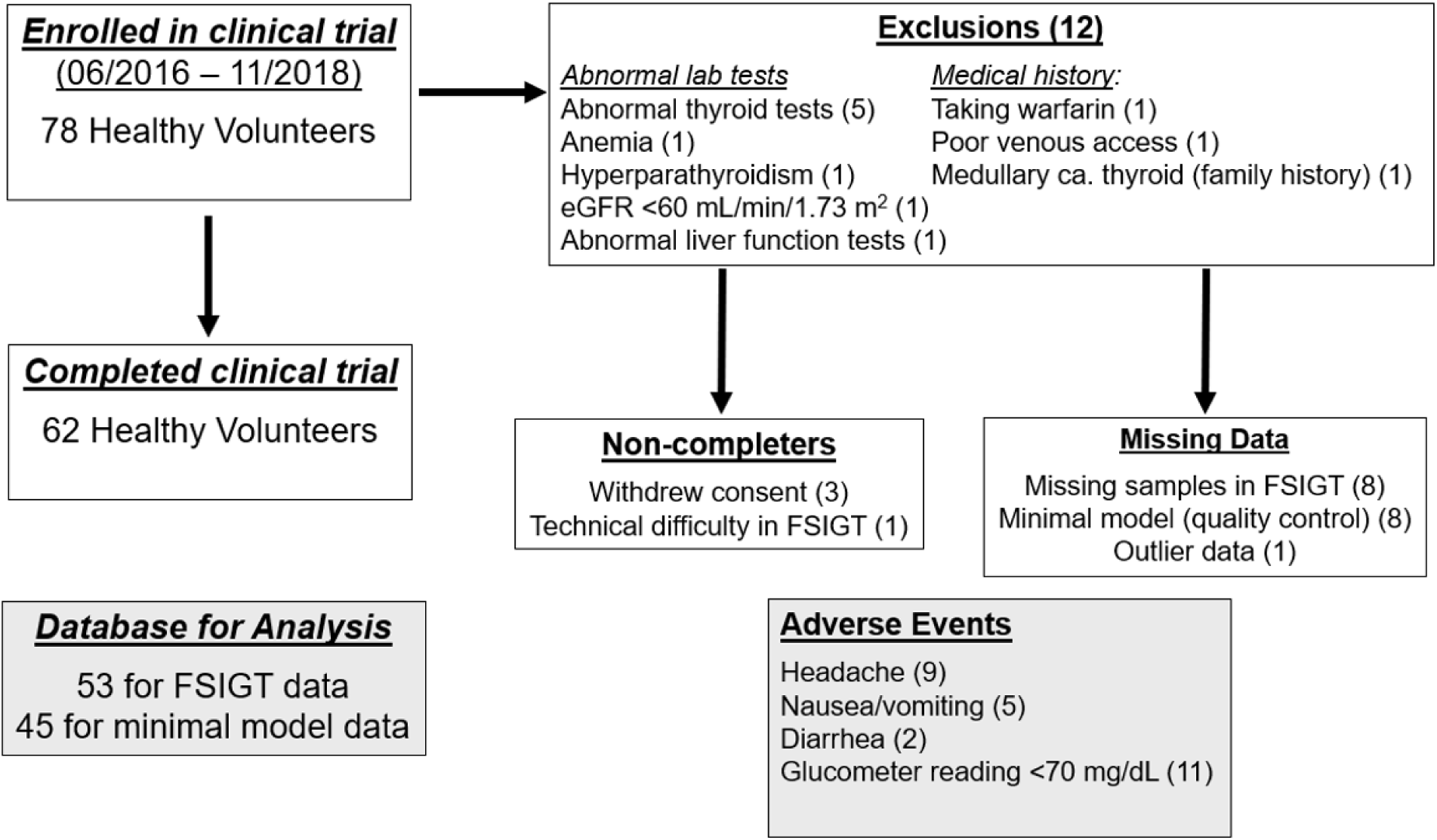
CONSORT diagram summarizing disposition of research participants. Seventy-eight individuals were enrolled in this clinical trial between June, 2016 – November, 2018. Sixty-two participants completed frequently sampled glucose tolerance tests. Twelve enrollees were excluded for the following reasons (Fig. S1): low hematocrit (N=1), abnormal TSH levels (N=5), abnormal liver function tests (N=1), family history of medullary carcinoma of the thyroid (N=1), primary hyperparathyroidism (N=1), low eGFR (N=1), poor venous access (N=1), and treatment with warfarin (N=1). Thus, 66 individuals were judged to be eligible for the clinical trial. Three individuals changed their minds and withdrew from the study after providing informed consent but before undergoing an intravenous glucose tolerance test; technical challenges during one FSIGT led to withdrawal of an additional participant. The remaining 63 participants completed the study. Ten individuals were excluded from the data base either because of missing data (N=9) or outlier data (N=1). Thus, data from 53 individuals comprised the final database for most of the analyses. Data from eight individuals’ placebo FSIGTs could not be analyzed with the Minimal Model software because of issues related to data quality. Accordingly, Minimal Model analysis of the FSIGT was based on data from 45 individuals. There were 27 reported adverse events of mild to moderate severity. Two participants reported feeling light-headed or dizzy in the evening after leaving the Amish Research Clinic; these were classified as probably being related to the study. There were 11 events in which a participant’s plasma glucose (determined using a glucometer) was <70 mg/dL (52-63 mg/dL) at the end of an FSIGT. Three participants reported symptoms (weakness, lightheadedness, or not feeling well) 60-90 minutes after receiving intravenous glucose, at which time glucometer readings of 42 – 60 mg/dL were obtained. Symptoms resolved quickly in all cases without intervention, and the FSIGT was not terminated. These were judged to be mild in severity and probably caused by the study.

**Figure S2.**
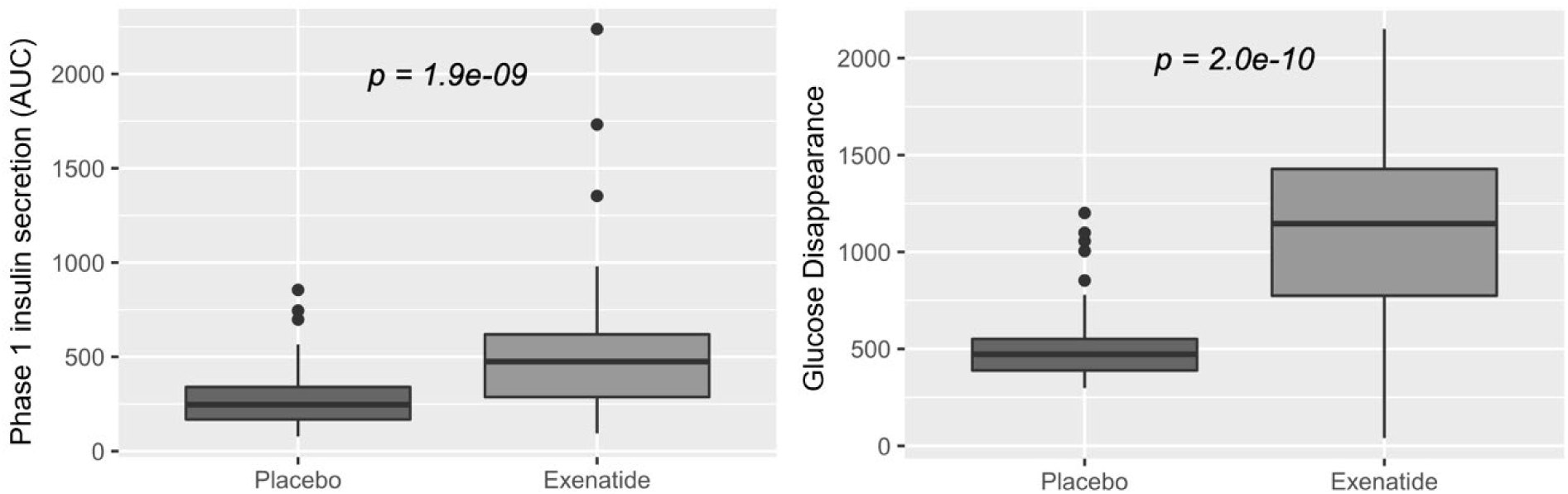
Exenatide increases first phase insulin secretion and the rate of glucose disappearance in frequently sampled intravenous glucose tolerance tests. Data are presented as means of data for 53 research participants. The boxes encompass the 2^nd^ and 3^rd^ quartiles; the “whiskers” correspond to the top and bottom quartiles (excluding outliers). “Outliers” are depicted by the closed circles. Statistical significance (calculated using a paired t-test) is indicated on the graphs. Areas under the curve for 1^st^ and 2^nd^ phase insulin secretion are expressed in the following units: (µU/mL) · min.

**Figure S3.**
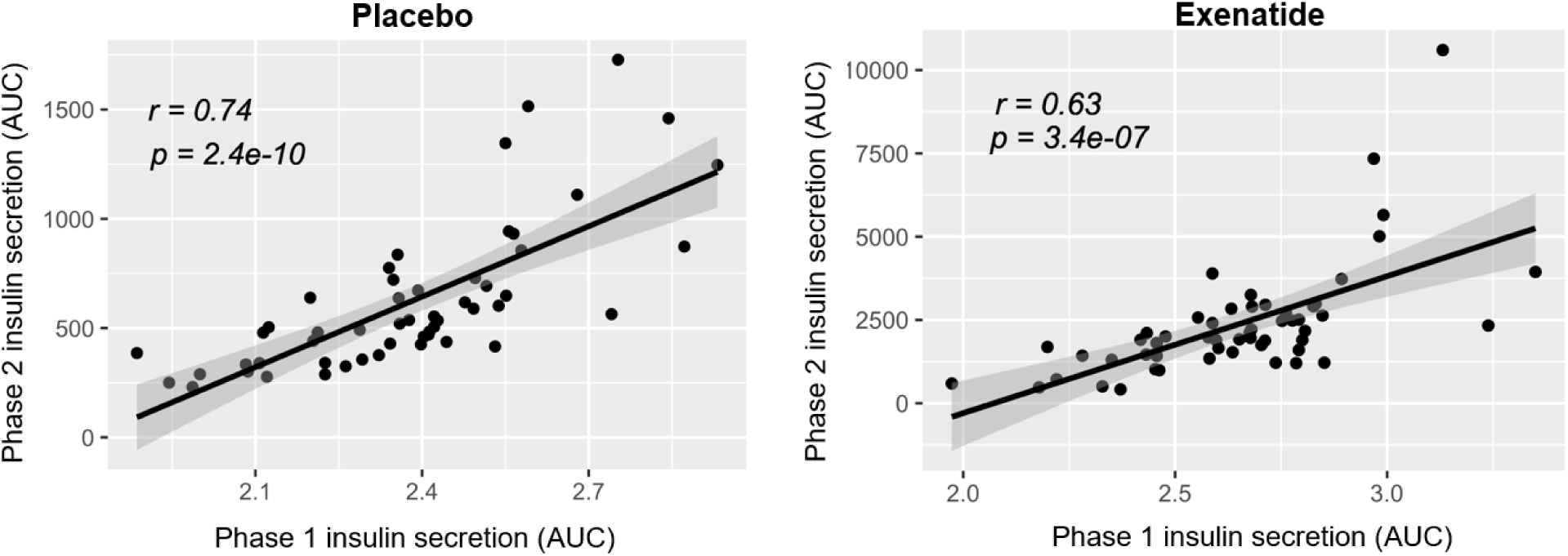
Correlations between first and second phase insulin secretion. Intravenous glucose triggers a biphasic response in the FSIGT: first phase insulin secretion (0-10 min) and second phase insulin secretion (10-50 min). Data are plotted as the logarithms of the areas under the curves for first and second phase insulin secretion; placebo studies are plotted in the left panel and exenatide studies in the right panel. Values for correlation coefficients (r) and p-values are indicated in the figure. Areas under the curve for 1^st^ and 2^nd^ phase insulin secretion are expressed in the following units: (µU/mL) · min.

**Figure S4.**
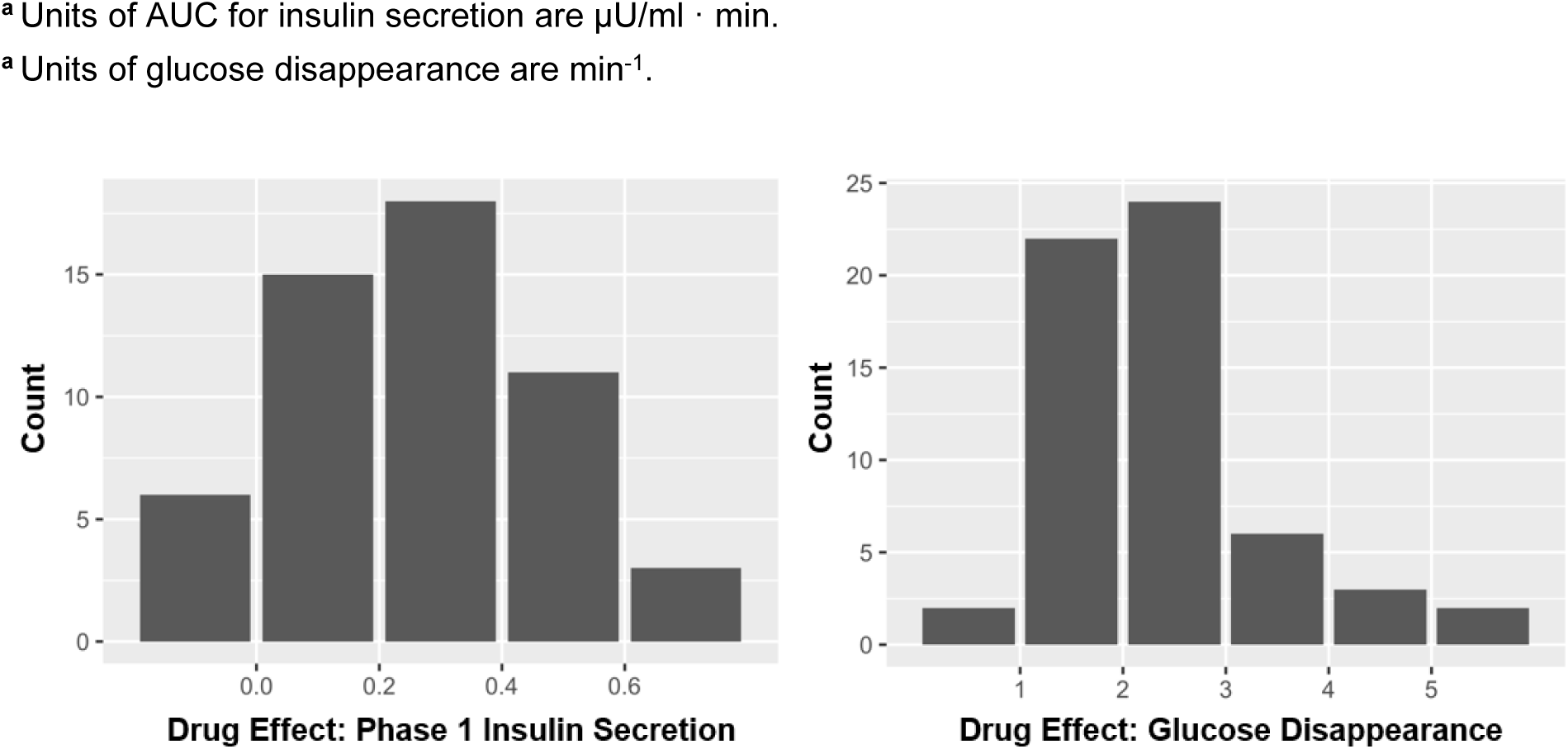
Inter-individual variation in magnitude of responses to exenatide: first phase insulin secretion and rate of glucose disappearance. Indices for effects of exenatide on first phase insulin secretion**^a^** and the rate of glucose disappearance**^b^** are defined in Table 1. Histograms depicting inter-individual variation are presented in the left panel for first phase insulin secretion and the right panel for the rate of glucose disappearance.

